# Pulmonary Distribution and Lesion Penetration of Antimicrobials in Patients with Nontuberculous Mycobacterial Disease

**DOI:** 10.64898/2026.05.12.26352725

**Authors:** Fumiya Watanabe, Miyako Hiramatsu, Toru Kawakami, Teruaki Oka, Hayato Nanami, Kiyomi Shimoda, Kazuhiko Hanada, Yuji Shiraishi, Kozo Morimoto

## Abstract

**Background:** The intrapulmonary pharmacokinetics of antimicrobial agents used to treat nontuberculous mycobacterial (NTM) pulmonary disease remain poorly characterized, limiting the optimization of dosing regimens. This study characterized the plasma and intrapulmonary pharmacokinetics of azithromycin, ethambutol, rifampicin, clofazimine, and amikacin, as well as their penetration into pulmonary lesion sites.

**Methods:** We prospectively enrolled patients undergoing guideline-based treatment for NTM pulmonary disease who were indicated for surgical resection at a single center in Japan. Drug concentrations were measured in the plasma and lung samples, and analyzed using a population pharmacokinetic model. The lung lesion site, cavity, or nodule/bronchiectatic were evaluated as covariates of the plasma-to-lung partition ratios.

**Results:** Twenty-four patients were enrolled in the study. Antimicrobial agents other than rifampicin and amikacin accumulate in the lungs at concentrations > 40-fold higher than those in the plasma. Notably, the intrapulmonary half-life of ethambutol, which has not been well-characterized to date, is estimated to be approximately 2 months, indicating prolonged retention within the lungs. Evaluation of drug penetration into cavities and nodular/bronchiectatic lesions showed no clearly reduced concentration compared to that of normal lung tissue. However, in the single case where the caseum was obtained, azithromycin, ethambutol, and rifampicin levels exhibited clearly lower concentrations.

**Conclusions:** Ethambutol shows a prolonged intrapulmonary half-life, suggesting sustained lung exposure even with intermittent dosing. The absence of clearly reduced drug penetration into lesion sites suggests that lesion phenotype alone may have limited value in guiding drug selection.

**Key points:** Ethambutol exhibited a prolonged intrapulmonary half-life of 2 months. Clofazimine
continued to accumulate in the lung for at least 3 years. Evaluation of drug penetration into pulmonary
lesions showed no clearly reduced concentration compared to that of normal tissue.

## INTRODUCTION

The incidence and prevalence of nontuberculous mycobacterial (NTM) pulmonary disease are increasing worldwide, with *Mycobacterium avium* and *M. abscessus* accounting for most infections [1, 2]. NTM pulmonary disease involves multiple types of pulmonary lesions, including nodular bronchiectatic or fibrocavitary forms [1, 3], which frequently coexist and various clinical manifestations. From these, cavitation and severe bronchiectasis are generally associated with a poor prognosis [4], requiring the addition of parenteral amikacin to oral antimicrobial therapy [1]. More recently, liposomal amikacin prepared for inhalation has been approved for patients with *M. avium* complex (MAC) pulmonary disease who fail to achieve culture conversion after at least 6 months of standard therapy [5]. This highlighted the increasing importance of antimicrobial selection and dose optimization according to antimicrobial penetration characteristics into disease phenotype.

Drug dose optimization is typically guided by pharmacokinetic-pharmacodynamic analysis based on plasma drug concentrations; however, plasma concentrations do not necessarily correlate with intrapulmonary concentrations [6]. As anti-NTM pulmonary disease drugs are needed to act on the lung, intrapulmonary pharmacokinetics are important. However, as sampling from the lungs is more invasive and complex than plasma sampling, intrapulmonary pharmacokinetic data is limited. One study quantified concentration of first-line anti-tuberculosis agents in the epithelial lining fluid and alveolar cells in the lungs of 157 patients with tuberculosis [7]. Although rifampicin did not show marked accumulation in the lungs relative to the plasma, ethambutol achieved substantially higher concentrations. However, this study evaluated only the lung-to-plasma concentration ratio of ethambutol and did not investigate parameters such as the intrapulmonary half-life. Since ethambutol is an important drug for preventing macrolide resistance in MAC pulmonary disease [8, 9], elucidating its intrapulmonary pharmacokinetics could directly inform dosing optimization.

Another study has demonstrated that lesion type and heterogeneity influence drug penetration into lesion [10]. Moreover, it has been suggested that the extent of lesion penetration affects the sterilizing efficacy against mycobacterial populations there by animal infection model [11–13]. Strydom et al. [10] investigated the potential of lesion penetration of seven major antituberculosis drugs in 15 patients, finding that rifampicin and clofazimine exhibited comparable penetration into cavity wall relative to normal lung tissue. Notably, clofazimine shows low penetration into the large necrotic granuloma. However, neither study examined the penetration of azithromycin and amikacin, key drugs for NTM pulmonary disease, into lung lesions.

While intrapulmonary pharmacokinetics are relatively well-characterized for anti-tuberculosis drugs, such information remains very limited for anti-NTM pulmonary disease drugs, and further evidence is needed to optimize their use. Furthermore, improved intrapulmonary pharmacokinetic data may help inform patient selection for inhaled vs intravenous amikacin therapy. Therefore, this study characterized the intrapulmonary pharmacokinetics of antimicrobials used for NTM pulmonary disease, including inhaled amikacin, as well as distributions in pulmonary lesions.

## METHODS

### Study Design

This prospective, single-center, observational study was conducted at Fukujuji Hospital, Tokyo, Japan. This study was conducted in accordance with the ethical principles of Declaration of Helsinki. The Institutional Review Boards of Fukujuji Hospital (no. 23018) and Meiji Pharmaceutical University (no. R5-004) approved the study. All participants provided written informed consent.

### Patient Population

Patients aged ≥ 20 years who underwent guideline-based treatment for *M. avium* or *M. abscessus* pulmonary disease between November 2023 and February 2026 and were indicated for surgical intervention were enrolled in this study. Patients for whom dosing or administration times were unknown were excluded from this study. Considering the limited number of surgical cases of NTM pulmonary disease, sample size was not prespecified, and all patients enrolled during the study period were included in the analysis.

### Pharmacokinetic Sampling

Antimicrobial agents were administered either on the morning of surgery or evening prior to surgery, and the administration times were recorded. Doses were administered according to guideline-recommended regimens [1]: azithromycin 250 mg/day, ethambutol 10–15 mg/kg/day, rifampicin 10 mg/kg/day, clofazimine 100 mg/day, intravenous amikacin 10–15 mg/kg/day, and inhaled amikacin 600 mg/day. Owing to constraints of the Japanese healthcare reimbursement system, amikacin was administered as a nebulized intravenous formulation rather than an amikacin liposome inhalation suspension.

Blood samples were collected at the start and end of surgery. These were deproteinized using acetonitrile or 10% trichloroacetic acid and drug concentrations were quantified using a validated high-performance liquid chromatography-mass spectrometry method [14].

Resected lung tissues were briefly rinsed with saline and subsequently dissected under a microscope to discriminate between normal lung tissue and lesion sites (cavitary and nodular/bronchiectatic lesions). To characterize intra-individual variability in drug distribution within normal lung tissue, normal lung parenchyma was sampled from four regions surrounding the lesion (superior, inferior, and bilateral) and each sample was analyzed separately. The lung tissue samples were homogenized, deproteinized, then quantified [14].

The lower limits of quantification were 0.01 μg/mL for azithromycin, 0.02 μg/mL for ethambutol and clofazimine, 0.05 μg/mL for rifampicin, and 5 μg/mL for amikacin.

### Penetration Ratios into Lung Lesions

For each patient, the average across the four normal lung tissue samples was calculated and used as the reference value. The lesion-to-normal concentration ratio was then computed for each drug and lesion type to determine pulmonary penetration.

### Pharmacokinetic Modeling

Population pharmacokinetic models were developed using NONMEM (version 7.6.0; ICON Development Solutions, Hanover, MD, USA), following a two-stage approach. First, plasma-only pharmacokinetic models were developed independently for each drug. For intravenous amikacin, therapeutic drug monitoring data obtained within 1 week prior to surgery were also included in the model building. In the second stage, plasma and intrapulmonary pharmacokinetic data were analyzed using a tissue penetration model.

For the plasma models, one- and two-compartment models were tested. Since plasma concentration data during the absorption phase were unavailable, absorption-related pharmacokinetic parameters (absorption rate constant and lag time) could not be estimated and were fixed at values reported in the literature [15–19]. Considering the sparse sampling design (two to three plasma measurements per individual), error was modeled using either an additive or proportional strategy, and inter-individual variability was estimated for clearance only. The base model was selected using the Akaike Information criterion value, supported by relative standard error values and goodness-of-fit plots. Stepwise covariate modeling [20] was applied to identify significant covariates for the model (co-administration of rifampicin for azithromycin clearance and renal function assessed by Cockcroft–Gault estimated creatinine clearance or estimated glomerular filtration rate for ethambutol and amikacin clearance).

The penetration of each drug into the lungs was determined using a separate lesion compartment, as described by Anderson et al. [21], according to the following equations:

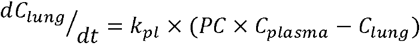

where *C* denotes drug concentration, *k*_*p*l_ is the plasma-lung equilibration rate constant (/h), and *PC* is the partition coefficient representing the equilibrium ratio of lung-to-plasma drug concentration. Cavitary and nodular/bronchiectatic lesions were evaluated as candidate covariates for PC using a likelihood ratio test at a significance level of 5% (corresponding to a decrease in the objective function value ≥ 3.84).

## RESULTS

Overall, 24 patients were enrolled and their characteristics at surgery are summarized in Table 1. Reflecting the surgical indication of patients enrolled, a high proportion presented cavitary disease (67%) and macrolide resistance (42%). The most frequently resected sites were the right middle lobe and left lingular segment. Of 24, 19 patients (79%) received ethambutol. Detailed information on concomitant antimicrobial therapy, surgical sites, and resected lesion types for each patient is provided in Table S1.

**Table 1.** Baseline characterization of study participants.

| Characteristics | Value |
| --- | --- |
| Age (year) | 60 ± 7.0 |
| Female | 22 (88%) |
| Body weight (kg) | 52 ± 6.8 |
| Bosy mass index (kg/m<sup>2</sup>) | 20 ± 2.0 |
| Causative bacterium |  |
| *M. avium* | 21 (88%) |
| *M. abscessus* | 3 (13%) |
| Radiological findings |  |
| Noncavitary-nodular bronchiectatic form | 8 (33%) |
| Cavitary- nodular bronchiectatic form | 14 (58%) |
| Fibro-cavitary form | 2 (8.3%) |
| Macrolide resistant | 10 (42%) |
| Follow-up term (months) | 28 [12–38] |
| Serum creatinine level (mg/dL) | 0.71 ± 0.12 |
| estimated glomerular filtration rate (mL/min/1.73m<sup>2</sup>) | 69 ± 11 |
| Creatinine clearance (mL/min) | 73 ± 18 |
| Mean ± standard deviation, Median (interquartile range), or Count (%) |  |

Measured drug concentrations in plasma, normal lung tissue, and lesion sites are shown in Figure 1. Two concentration measurements, one for rifampicin and other for amikacin, fell below the lower limit of quantification and were excluded from subsequent analyses. Azithromycin and clofazimine achieved higher concentrations in the lung tissues than in the plasma. Ethambutol accumulated substantially in the lungs, with concentration markedly elevated at 24 h post-administration.

**Figure 1.**
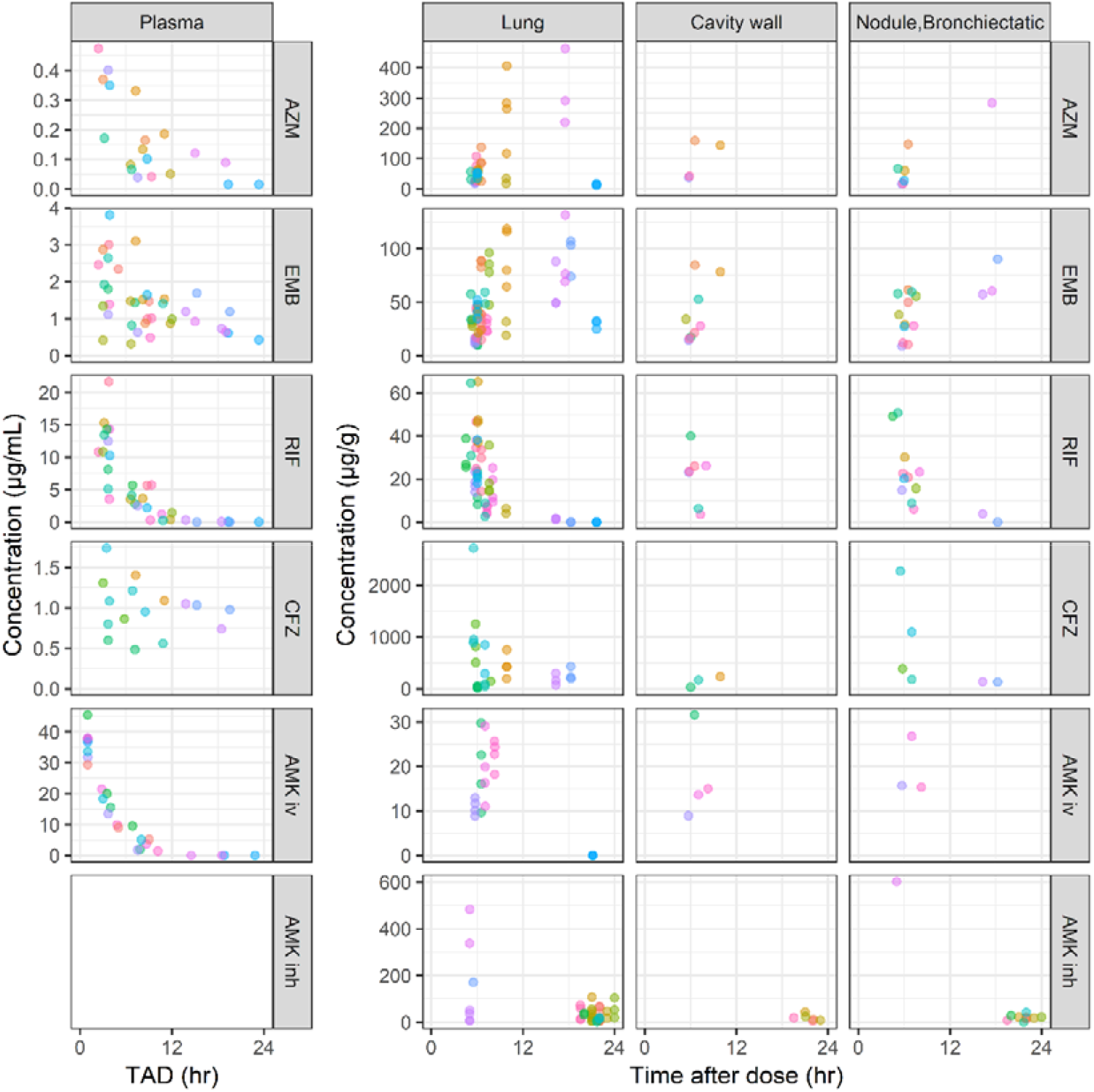
Concentration–time profiles of antimicrobial agents in plasma and lung tissues. Measured drug concentrations in plasma (μg/mL) and lung tissue (μg/g) are plotted against time after dosing for each patient (color-coded by individual). AZM, azithromycin; EMB, ethambutol; RIF, rifampicin; CFZ, clofazimine; AMK iv, intravenous amikacin; AMK inh, inhaled amikacin; TAD, time after dosing.

None of the drugs showed lower penetration into the lesion sites (Figure 2). However, drug concentrations in lung tissues exhibited substantial intra-individual variability, with coefficients of variation of 36%, 25%, 34%, 47%, 28%, and 60% for azithromycin, ethambutol, rifampicin, clofazimine, intravenous amikacin, and inhaled amikacin, respectively, which was reflected in the high variability in penetration into pulmonary lesion sites. One sample obtained from a cavitary wall was identified as caseum, and the concentrations of azithromycin, ethambutol, and rifampicin in this sample were markedly lower than those in normal lung tissue (Figure 2).

**Figure 2.**
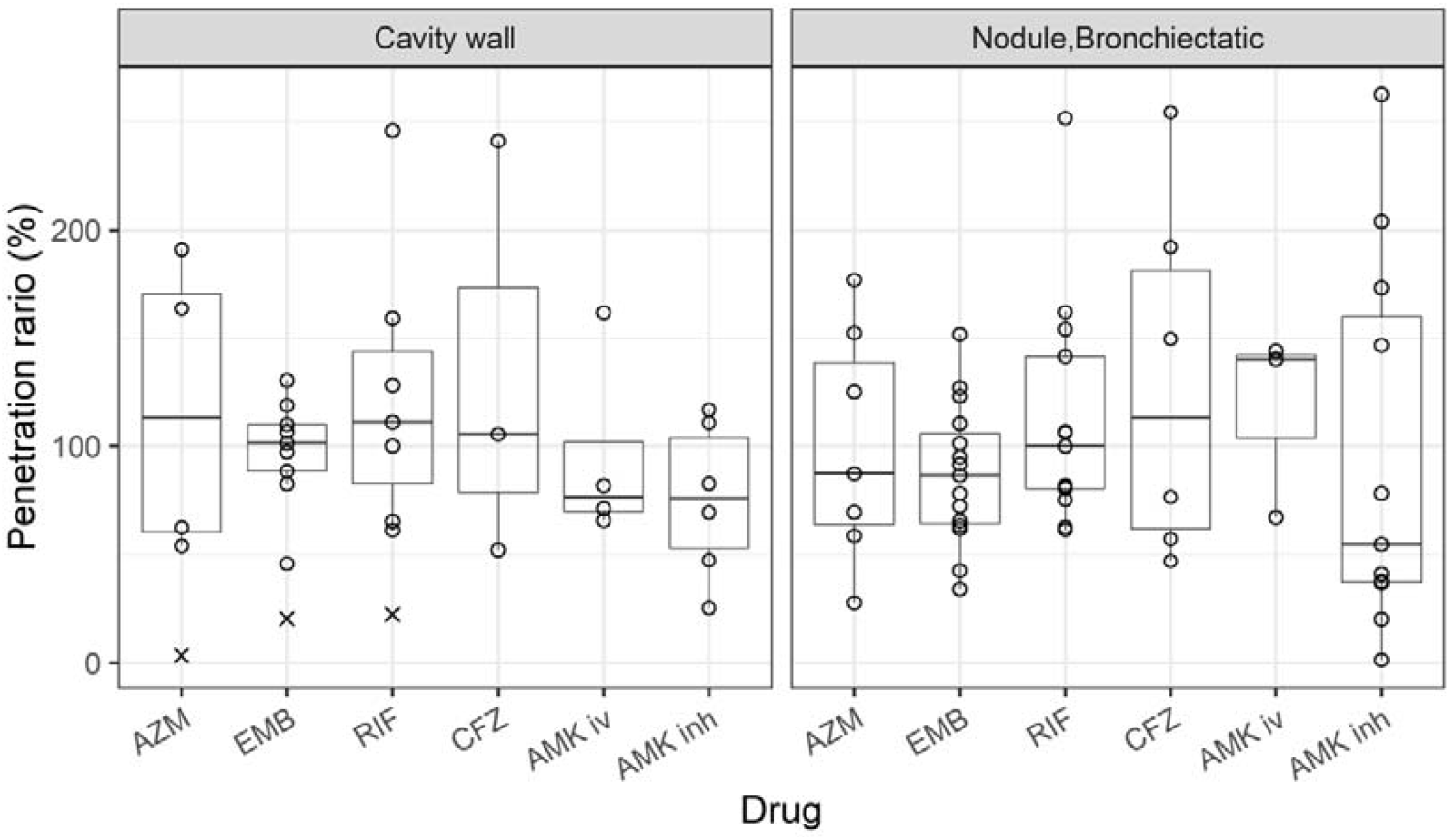
Drug penetration ratios into pulmonary lesions. Penetration ratio was calculated as the ratio of the lung lesion concentration divided the mean concentration in normal lung tissue of each patient, expressed as a percentage. A value of 100% indicates penetration equivalent to that of normal lung tissue. Circles represent individual values, whereas crosses (×) represent the values obtained from the single caseum sample. AZM, azithromycin; EMB, ethambutol; RIF, rifampicin; CFZ, clofazimine; AMK, intravenous amikacin; AMK, inhaled amikacin.

Pharmacokinetic modeling was performed to estimate intrapulmonary pharmacokinetic parameters, such as intrapulmonary half-life, relevant to dosing optimization. For the plasma models, a two-compartment model best described the concentrations profiles of azithromycin and ethambutol, whereas a one-compartment model provided the best fit for the other drugs. All tested covariate candidates were incorporated in the final models. In lung pharmacokinetic models, lesion type was evaluated as a candidate covariate for pulmonary partition coefficients; however, neither cavitary nor nodular/bronchiectatic lesions achieved significance. The goodness-of-fit plots (Figures S1 and S2) and visual predictive checks (Figure 3) demonstrated the robust predictiveness of the model. The model-predicted plasma-to-lung partition coefficients and intrapulmonary half-lives were of 535-fold and 6.3 months for azithromycin, 40-fold and 2.0 months for ethambutol, 5.2-fold and 6.3 min for rifampicin, 18800-fold and 22 years for clofazimine, and 1.3-fold and 2.3 h for intravenous amikacin. The estimated intrapulmonary half-life of inhaled amikacin exceeded that of intravenous amikacin by approximately 5 h. The detailed final model structures and parameter estimates are listed in Tables S2–S6.

**Figure 3.**
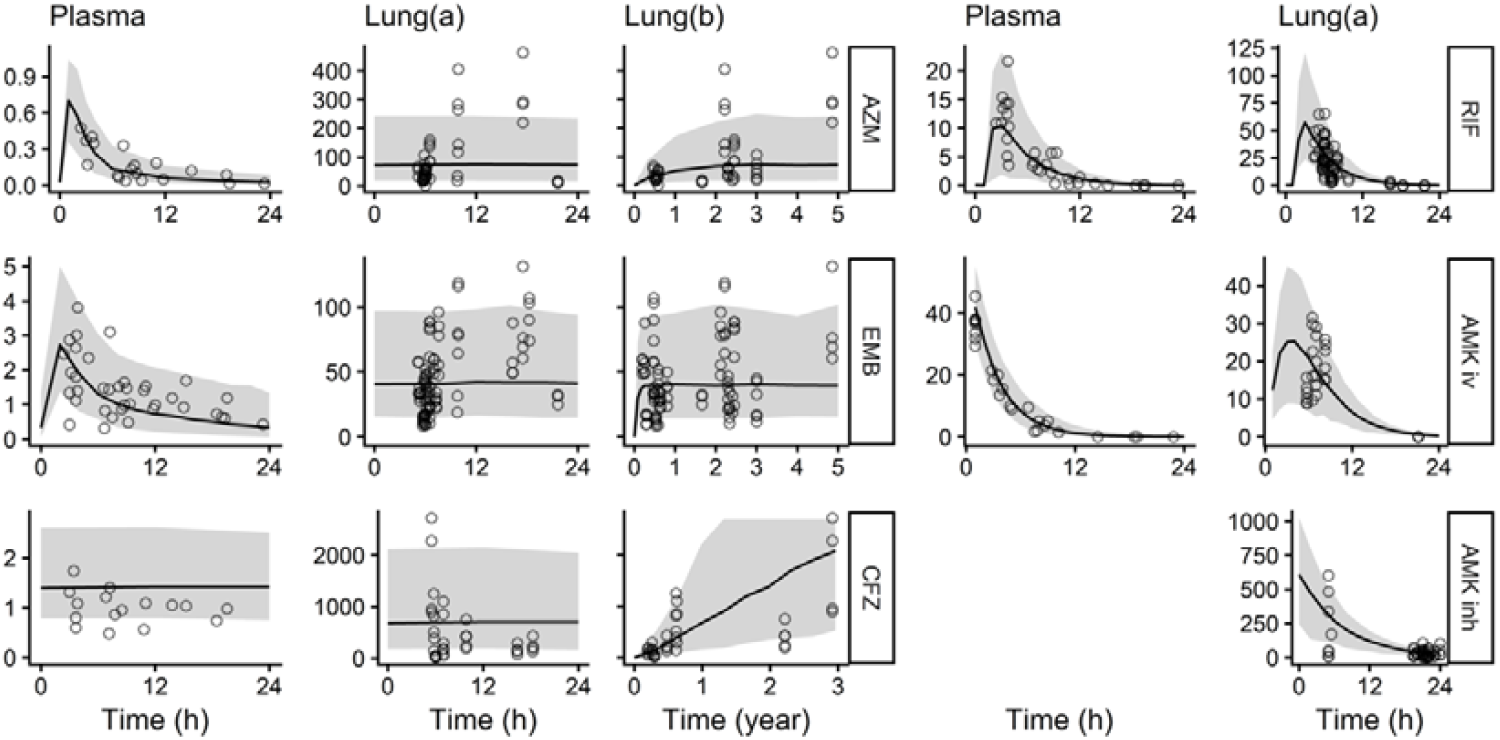
Model-predicted and measured drug concentration plots. Observed drug concentrations (circles) are plotted against model-predicted mean concentration–time profiles (solid lines), with shaded areas representing the 90% prediction interval. For all drugs, the y-axis represents measured concentration (μg/mL for plasma; μg/g for lung tissue) and x-axis represents time after the last dose (h). For azithromycin, ethambutol, and clofazimine, which exhibited marked pulmonary accumulation, additional plots of lung concentration versus time since the first dose (years) are presented (lung [b]) to capture the full accumulation trajectory. AZM, azithromycin; EMB, ethambutol; RIF, rifampicin; CFZ, clofazimine; AMK iv, intravenous amikacin; AMK inh, inhaled amikacin.

## DISCUSSION

This study provides the characterization of the intrapulmonary pharmacokinetics of key antimicrobials used in NTM pulmonary disease, with novel findings demonstrating the prolonged intrapulmonary half-life of ethambutol and clofazimine as well as first evaluation of the penetration of azithromycin, ethambutol, and amikacin in this clinical context.

Ethambutol demonstrated marked pulmonary accumulation, with a lung-to-plasma concentration approximately 40-fold higher than plasma and a rifampicin ratio of 5.2-fold. These findings are broadly consistent with those of McCallum et al. [7], who reported plasma-to-lung concentration ratios in the alveolar cells of 15-fold for ethambutol and 1.4-fold for rifampicin. The quantitatively higher ratios observed in this study likely reflect methodological differences, as homogenized lung tissue captures both intracellular and extracellular drug fractions, whereas isolated alveolar cell measurements do not. Ethambutol concentrations in the lung exhibited minimal intraday fluctuations, with an estimated intrapulmonary half-life of approximately 2 months, a novel finding with clinical importance. Thus, prolonged retention even with intermittent dosing, similar to azithromycin, may provide a mechanistic explanation for t suppression of macrolide resistance in *M. avium* pulmonary disease, even under a three-times-weekly regimen [22].

This is the first study to evaluate the intrapulmonary concentrations of clofazimine during long-term treatment. Clofazimine was orally administered for up to 3 years, whereby time its concentration in the lungs continued to increase without reaching a plateau. Therefore, the relative standard error of lung half-life (22 years) estimated using the population pharmacokinetic model was 706%, suggesting an inherent imprecision of estimating this parameter (Table S5). Clofazimine is highly lipophilic and is known to accumulate at high concentrations in various organs, with tissue deposition of crystallized clofazimine reported particularly during high-dose (≥ 300 mg/day) or long-term administration [23], which may explain the accumulation observed in this study.

Inhaled amikacin achieved a mean lung concentration of 26.9 μg/g (standard deviation, 24.7) at approximately 20 h post-inhalation, broadly compared to peak lung concentrations following intravenous administration. Siegel et al. [24] and Kurahara et al. [25] reported amikacin resistance rates of 18% and 15.9%, respectively, among patients with NTM pulmonary disease treated with amikacin liposome inhalation suspension. Sustained drug exposure in the lungs, while therapeutically desirable, may simultaneously contribute to resistance. However, as the inhaled amikacin used in this study was not a liposomal formulation, further investigation is required to clarify this association.

Penetration into cavitary and nodular/bronchiectatic lesions, relative to normal lungs tissue was not markedly reduced for any of the tested drugs (Figure 2), consistent with finding by Strydom et al. [10] in tuberculosis, where rifampicin, clofazimine, and kanamycin exhibited penetration into the cavity wall comparable to normal lungs. Strydom et al. further demonstrated that clofazimine concentrations within the caseum were markedly lower than those in normal lungs, similar to what was observed in this study for azithromycin, ethambutol, and rifampicin, although in only one caseum (Figure 2). These findings may suggest that caseous lesions are less responsive to medical therapy.

This study had several limitations. First, the small sample size limited statistical power to detect differences in lesion penetration and precluded adequate characterization of inter-individual variability in pulmonary drug concentrations. Second, the analysis was based on limited sampling time points, resulting in low precision of several population parameter estimates, which was evident for clofazimine, where lung concentrations did not plateau during the observation period. This resulted in relative standard errors of 676% and 706% for the lung partition coefficient and intrapulmonary half-life, respectively. Therefore, the temporal changes in tissue distribution during long-term therapy cannot be fully characterized, although the observation that clofazimine continues to accumulate in the lungs for at least 3 years is itself clinically relevant. Third, a direct relationship between the plasma and pulmonary concentrations, and clinical outcomes, including treatment response and resistance suppression, has not been fully established. Finally, the potential influence of concomitant medications and patient-specific factors (e.g., disease severity and inflammatory status) could not be fully controlled. Additionally, residual confounding may have contributed to the observed variability in pulmonary drug concentrations.

This study demonstrates that ethambutol exhibits a prolonged half-life in the lungs and persists within pulmonary tissues, supporting the use of intermittent dosing intervals and providing mechanistic evidence for its role in preventing macrolide resistance. However, the dose required to effectively suppress resistance remains unclear, and the present findings are informing dosing intervals rather than defining optimal dosing levels. Furthermore, none of the drugs clearly demonstrated reduced penetration into the lesion sites, except in the caseous necrotic regions. Thus, lesion phenotype alone may have limited utility in guiding drug selection, although the rarity of caseum sampling in this study warrants caution in generalizing this conclusion.

## Notes

### Author Contributions

F.W. and Y.S. conceptualized the study. F. W. and M. H. curated the data. M.H., T.K., H.N., K.S., and Y.S. performed radiological assessments of the patients. T.O. made the pathological diagnosis and dissected the samples according to the lesion sites. F.W. and K.H. performed formal analyses. K.H. provided the software for the analysis. M.H., Y.S., and K.M. supervised and performed the experiments. M.H., T.K., H.N., K.S., and Y.S. provided informed consent. F.W., K.H., and K.M. prepared original drafts of the manuscript. M.H., T.K., T.O., H.N., K.S., K.H., Y.S., and K.M. reviewed, edited and approved the manuscript.

### Conflict of Interest

The authors declare no conflicts of interest.

### Funding

The Japan Research Foundation for Clinical Pharmacology 2023 (F.W.) supported this work.

## Acknowledgments

We thank Editage (www.editage.jp) for the English language editing.

## Declaration of generative AI and AI-assisted technologies in the manuscript preparation process

No generative AI or AI-assisted technologies were used to prepare this manuscript.

## Ethics Statement

This study was approved by the institutional review boards of Fukujuji Hospital (no. 23018) and Meiji Pharmaceutical University (no. R5-004). All participants provided written informed consent.

## Data Availability Statement

The data supporting the findings of this study are available from the corresponding author upon reasonable request.

## Notes

### Competing Interest Statement

The authors have declared no competing interest.

### Author Declarations

The Institutional Review Boards of Fukujuji Hospital (no. 23018) and Meiji Pharmaceutical University (no. R5-004) approved the study.

