## Supplement for "Pulmonary Distribution and Lesion Penetration of Antimicrobials in Patients with Nontuberculous Mycobacterial Disease"

### **Table S1. Demographic information of the study population.**

| ID | Gender | Age  (year) | BMI  (kg/m^2^) | Measured Drugs | Resected lesion types | Surgical sites |
| --- | --- | --- | --- | --- | --- | --- |
| 1 | Male | 50’s | 20.1 | EMB, RIF, CFZ, AMK iv | Cavity | RUML |
| 2 | Female | 60’s | 21.6 | AZM, EMB, RIF, AMK iv | Nodule, Cavity | RUL |
| 3 | Female | 70’s | 17.2 | RIF, AMK iv | Nodule, Cavity | RUML |
| 4 | Female | 60’s | 17.2 | EMB, RIF, AMK iv | Nodule, Cavity | Lingula + S6 |
| 5 | Female | 40’s | 18.9 | AZM, EMB, RIF, AMK inh | Nodule, Cavity | RML + S2,6 |
| 6 | Female | 50’s | 23.6 | EMB, RIF, AMK inh | Nodule, Cavity | Lingula |
| 7 | Female | 60’s | 20.8 | EMB, AMK iv, AMK inh | Bronchiectasis | RML |
| 8 | Female | 60’s | 20.6 | AZM, EMB, AMK inh | Nodule, Cavity | RML + S6 |
| 9 | Female | 40’s | 18.3 | AZM, EMB, CFZ | Cavity | Lingula |
| 10 | Female | 50’s | 22.9 | AZM, EMB, RIF, AMK inh | Nodule, Cavity | Lingula |
| 11 | Female | 60’s | 21.6 | AZM, EMB, RIF, AMK inh | Nodule, Cavity | RML |
| 12 | Male | 50’s | 21.4 | EMB, AMK inh | Nodule, Cavity | Lingula + S3 |
| 13 | Male | 70’s | 19.1 | EMB, RIF, AMK inh | Nodule | RUL |
| 14 | Female | 50’s | 20.6 | CFZ | Nodule | RLL |
| 15 | Female | 60’s | 16.0 | RIF, AMK iv, AMK inh | Nodule | Right S6 |
| 16 | Female | 60’s | 20.1 | AZM, EMB, RIF, AMK inh | Nodule | RUL |
| 17 | Female | 50’s | 18.5 | EMB, RIF, CFZ | Nodule, Cavity | RML + S6 |
| 18 | Female | 50’s | 20.0 | CFZ, AMK inh | Nodule | Lingula |
| 19 | Female | 60’s | 18.0 | CFZ, AMK iv, AMK inh | Nodule | Lingula + S1,2 |
| 20 | Female | 60’s | 22.1 | AZM, EMB, RIF | Nodule, Caseum | RML + S6 |
| 21 | Female | 50’s | 23.3 | AZM, EMB, RIF, AMK iv | Not available | Lingula |
| 22 | Female | 50’s | 19.1 | EMB, RIF, CFZ, AMK inh | Nodule | Lingula + S8 |
| 23 | Female | 60’s | 19.4 | EMB, RIF, CFZ, AMK inh | Nodule | Lingula |
| 24 | Female | 50’s | 22.3 | AZM, EMB, AMK iv, AMK inh | Nodule | RML + S2 |

AZM, azithromycin; EMB, ethambutol; RIF, rifampicin; CFZ, clofazimine; AMK iv, intravenous amikacin; AMK inh, inhaled amikacin; RUL, right upper lobe; RUML, right upper and middle lobes; RML, right middle lobe; RLL, right lower lobe.

Segment numbers indicate that the resected segments were ipsilateral to the specified lobe. For instance, “RML + S6” denotes combined resection of the right middle lobe and the right S6 segment.

### **Table S2. Population pharmacokinetic parameters estimate for azithromycin.**

| Parameter (apparent) | Typical value (%RSE) | 95% CI |
| --- | --- | --- |
| Clearance (L/h) | 49 (13) | 36–65 |
| Volume of distribution (L) | 279 (31) | 147–578 |
| Inter-compartmental clearance (L/h) | 38 |  |
| Peripheral volume of distribution (L) | 355 |  |
| Absorption rate constant (/h) | 1.6 |  |
| Covariance | 1.9 (11) | 1.5–2.5 |
| BSV of clearance (%CV) | 13 (120) | 2.8–22 |
| Residuals _Plasma_ (%CV) | 56 (31) | 37–65 |
| PC (fold) | 535 (55) | 309–2940 |
| K _pl_ (/h) | 0.00015 (138) | 0.000010–0.067 |
| T _pl_ | 6.3 months |  |
| BSV of PC _lung_ (%CV) | 64 (49) | 24–82 |
| Residuals _Lung_ (%CV) | 66 (9.0) | 59–71 |

BSV, between-subject variability; CI, 95% confidence interval estimated using the bootstrap method (n = 1,000); CV, coefficient of variation; K_pl_, plasma-lung equilibration rate constant; PC, plasma-to-lung partition coefficient; T_pl_, equilibration half-life derived from ln (2)/K _pl_; RSE, relative standard error; 95%.

Covariate model: For clearance, rifampicin co-administration was incorporated as a binary covariate (0 or 1), expressed as CL_i_ = CL × 1.9 ^Rifampicin^, where CLi and CL are the individual and typical clearance values, respectively.

Residual error model: Residual error is expressed as C_i_ = C × (1 + ε), where C_i_ and C are individual observations and predictions, respectively, and ε is normally distributed with a mean of 0 and variance of σ^2^.

Parameters that could not be estimated, such as the absorption rate constant, were fixed at the values reported in the literature by Chotsiri et al. [1].

### **Table S3. Population pharmacokinetic parameters estimate for ethambutol.**

| Parameter (apparent) | Typical value (%RSE) | 95% CI |
| --- | --- | --- |
| Clearance (L/h) | 26 (12) | 20–34 |
| Volume of distribution (L) | 48 (46) | 19–144 |
| Inter-compartmental clearance (L/h) | 40 (26) | 23–69 |
| Peripheral volume of distribution (L) | 241 (43) | 112–1970 |
| Absorption rate constant (/h) | 0.44 |  |
| Absorption lag time (h) | 0.25 |  |
| Covariance | 1.4 (40) | 0.82–1.3 |
| BSV of clearance (%CV) | 42 (60) | 19–58 |
| Residuals _Plasma_ (%CV) | 50 (19) | 43–160 |
| PC (fold) | 40 (17) | 32–59 |
| K _pl_ (/h) | 0.00048 (110) | 0.00012–0.010 |
| T _pl_ | 2.0 months |  |
| BSV of PC _lung_ (%CV) | 50 (36) | 26–63 |
| Residuals _Lung_ (%CV) | 55 (10) | 49–60 |

BSV, between-subject variability; CI, 95% confidence interval estimated using the bootstrap method (n = 1,000); CV, coefficient of variation; K_pl_, plasma-lung equilibration rate constant; PC, plasma-to-lung partition coefficient; T_pl_, equilibration half-life derived from ln (2)/K _pl_; RSE, relative standard error; 95%.

For clearance, estimated glomerular filtration rate (eGFR, mL/min/1.73 m^2^) was included as a covariate, expressed as CL_i_ = CL × (eGFR / 69.5) ^1.4^, where CLi and CL are individual and typical values of clearance, respectively.

Parameters difficult to estimate, such as the absorption rate constant, were fixed at the values reported in the literature by Mehta et al [2].

### **Table S4. Population pharmacokinetic parameters estimate for rifampicin.**

| Parameter (apparent) | Typical value (%RSE) | 95% CI |
| --- | --- | --- |
| Clearance (L/h) | 9.8 (15) | 7.3–15 |
| Volume of distribution (L) | 4.4 (31) | 1.4–6.1 |
| Absorption rate constant (/h) | 0.25 |  |
| Absorption lag time (h) | 1.4 |  |
| BSV of clearance (%CV) | 31 (65) | 7.9–99 |
| Residuals _Plasma_ (%CV) | 71 (18) | 56–80 |
| PC (fold) | 5.2 (11) | 4.2–6.4 |
| K _pl_ (/h) | 6.6 (7) | 4.6–16 |
| T _pl_ | 6.3 minutes |  |
| BSV of PC _lung_ (%CV) | 38 (33) | 18–48 |
| Residuals _Lung_ (%CV) | 62 (8) | 57–68 |

BSV, between-subject variability; CI, 95% confidence interval estimated using the bootstrap method (n = 1,000); CV, coefficient of variation; K_pl_, plasma-lung equilibration rate constant; PC, plasma-to-lung partition coefficient; T_pl_, equilibration half-life derived from ln (2)/K _pl_; RSE, relative standard error; 95%.

Parameters that could not be estimated, such as the absorption rate constant, were fixed at the values reported by Nishimura et al [3].

### **Table S5. Population pharmacokinetic parameters estimate for clofazimine.**

| Parameter (apparent) | Typical value (%RSE) | 95% CI |
| --- | --- | --- |
| Clearance (L/h) | 2.9 |  |
| Volume of distribution (L) | 3360 |  |
| PC (fold) | 18800 (676) | 2000–1840000 |
| K _pl_ (/h) | 0.0000036 (706) | 0.000000036–0.000045 |
| T _pl_ | 22 years |  |
| BSV of PC _lung_ (%CV) | 60 (75) | 6.6–88 |
| Residuals _Lung_ (%CV) | 74 (5) | 70–77 |

BSV, between-subject variability; CI, 95% confidence interval estimated using the bootstrap method (n = 1,000); CV, coefficient of variation; K_pl_, plasma-lung equilibration rate constant; PC, plasma-to-lung partition coefficient; T_pl_, equilibration half-life derived from ln (2)/K _pl_; RSE, relative standard error; 95%.

Plasma pharmacokinetic parameters were obtained from the previously reported population pharmacokinetic model estimates from the same institution by Watanabe et al [4].

### **Table S6. Population pharmacokinetic parameters estimate for amikacin.**

| Parameter | Typical value (%RSE) | 95% CI |
| --- | --- | --- |
| Clearance (L/h) | 4.2 (4.7) | 3.8–4.6 |
| Volume of distribution (L) | 11 (6.0) | 9.6–12 |
| Covariance | 1 |  |
| BSV of clearance (%CV) | 15 (37) | 3.4–18 |
| Residuals _Plasma_ (%CV) | 41 (13) | 33–45 |
| PC (fold) | 1.3 (13) | 0.42–2.9 |
| K _pl_ (/h) for AMK inh | 0.14 (7) | 0.13–0.18 |
| K _pl_ (/h) for AMK iv | 0.3 |  |
| T _pl_ for AMK inh | 5.0 hours |  |
| T _pl_ for AMK iv | 2.3 hours |  |
| BSV of PC _lung_ (%CV) | 15 (91) | 9.3–225 |
| Residuals _Lung_ (%CV) | 88 (11) | 92–105 |

BSV, between-subject variability; CI, 95% confidence interval estimated using the bootstrap method (n = 1,000); CV, coefficient of variation; K_pl_, plasma-lung equilibration rate constant; PC, plasma-to-lung partition coefficient; T_pl_, equilibration half-life derived from ln (2)/K _pl_; RSE, relative standard error; 95%.

For plasma pharmacokinetics, creatinine clearance (CCR, mL/min) was included as a covariate of clearance, expressed as CL_i_ = CL × (CCR / 71.5) ^1^, where CLi and CL are individual and typical clearance values, respectively. Covariate effects were fixed to the estimates reported in the previous study by Watanabe et al. [5].

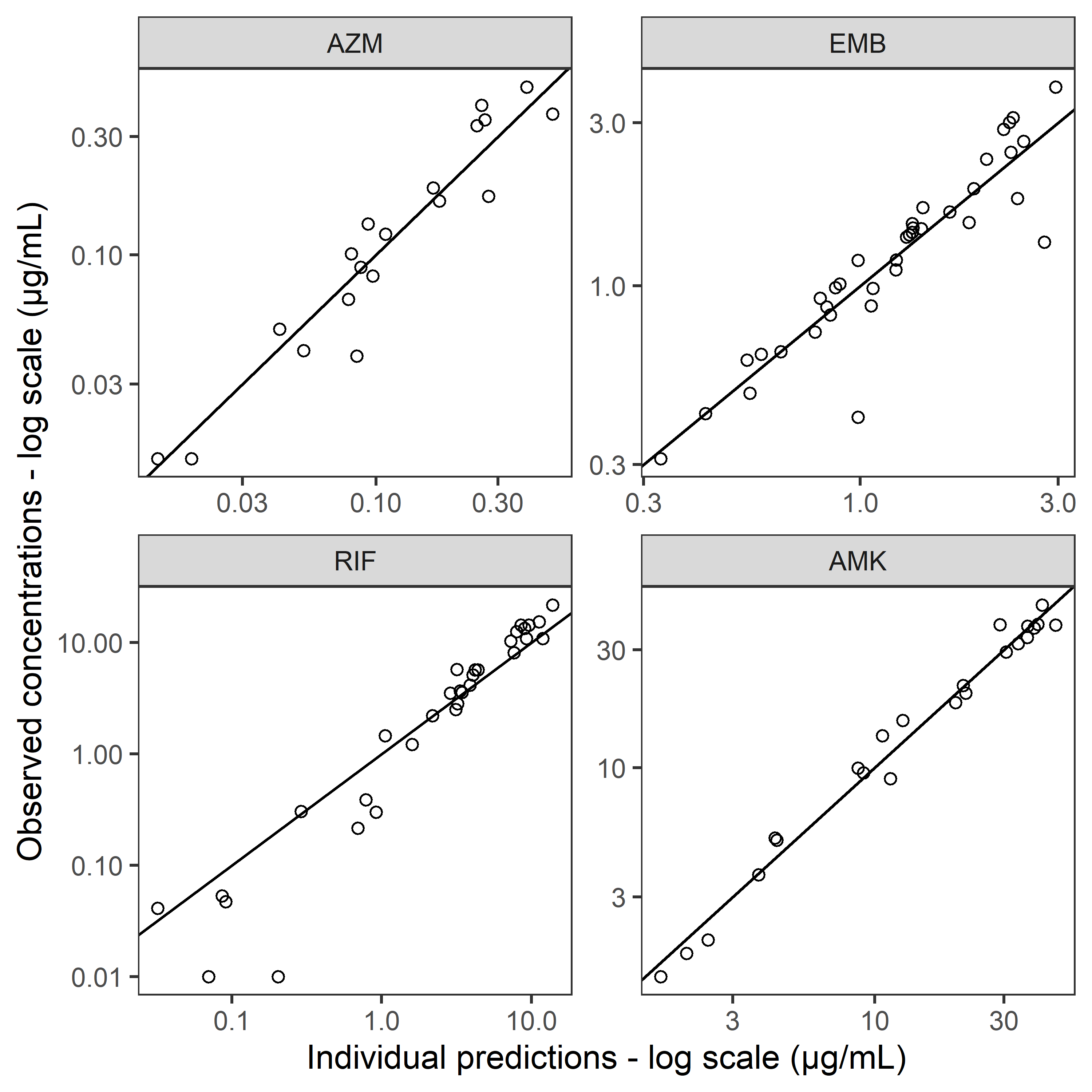

### **Figure S1. Goodness-of-fit plots for drug concentrations in plasma.**

Observed plasma concentrations plotted against individual model-predicted concentrations. The solid line represents line of identity. AZM, azithromycin; EMB, ethambutol; RIF, rifampicin; AMK, amikacin.

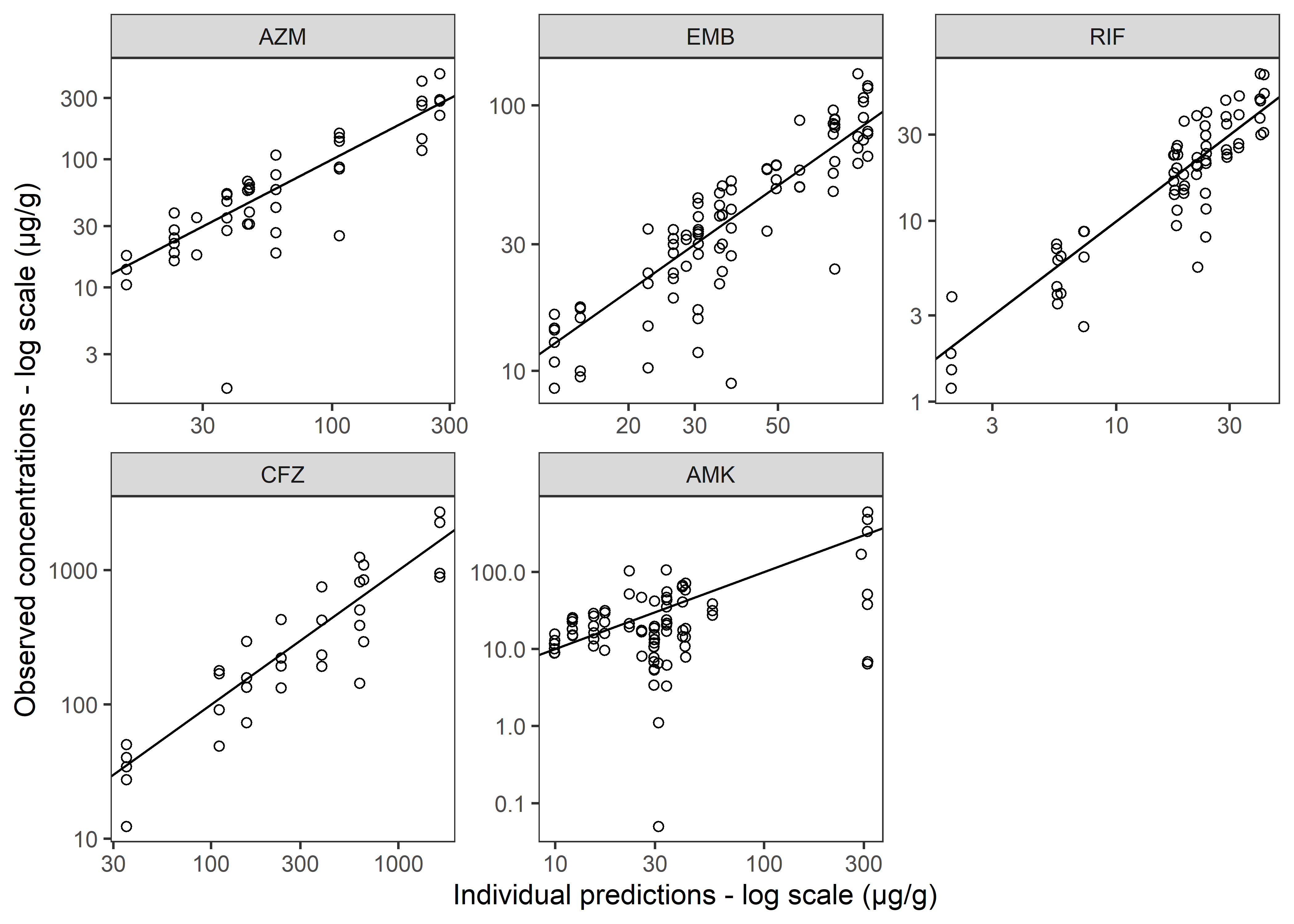

### **Figure S2. Goodness-of-fit plots for drug concentrations in the lung.**

Measured drug concentrations in the lung were plotted against individual model-predicted concentrations. Solid line, line of identity. AZM, azithromycin; EMB, ethambutol; RIF, rifampicin; CFZ, clofazimine; AMK, amikacin.
